# Estimating the effective reproduction number of the 2019-nCoV in China

**DOI:** 10.1101/2020.01.27.20018952

**Authors:** Zhidong Cao, Qingpeng Zhang, Xin Lu, Dirk Pfeiffer, Zhongwei Jia, Hongbing Song, Daniel Dajun Zeng

## Abstract

We estimate the effective reproduction number for 2019-nCoV based on the daily reported cases from China CDC. The results indicate that 2019-nCoV has a higher effective reproduction number than SARS with a comparable fatality rate.

**Article Summary Line:** This modeling study indicates that 2019-nCoV has a higher effective reproduction number than SARS with a comparable fatality rate.

## Text

As of 01/26/2020, the 2019 novel coronavirus (2019-nCoV), originated in Wuhan China, has spread to 29 mainland provinces, Hong Kong, Macau, Taiwan, as well as 11 other countries (*1, 2*). Early genome sequence and clinical studies of 2019-nCoV provided the evidence of human-to-human transmission and revealed its similarity to as well as differences from SARS (*3-5*). However, epidemiological investigations of 2019-nCoV are just beginning, and data-driven studies are critically needed to develop insights into this ongoing outbreak and evaluate the effectiveness of public health strategies, such as the currently implemented lockdown of Wuhan.

An important epidemiological understanding of 2019-nCoV is concerned with its transmissibility, quantified by the basic reproduction number *R*_0_ and the effective reproduction number *R. R*_0_ is the expected number of secondary infectious cases generated by an infectious case in a susceptible population. *R* is the expected number of secondary cases generated by an infectious case once an epidemic is underway (*6*). *R* = *R*_0_*x*, where *x* ∈ (0, 1) is the proportion of the population susceptible. Following (*7*), *R* is calculated as follows:

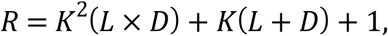

where *L* is the average latent period, *D* the average latent infectious period, *K* the logarithmic growth rate of the case counts as reported by China CDC. This form of *R* is appropriate because 2019-nCoV is still at its early growth stage. According to China CDC, we set *L* = 7 days and *D* = 9 days. Experiments with varying *L* and *D* values were also conducted.

Let *t* denote the number of days since the start of the outbreak and *Y*(*t*) the number of cases. *K* is estimated based on *Y*(*t*) at six time points. (Time-1) 12/31/2019, when the authorities reported the first 27 cases with the infection dated as early as 12/16/2019. As such, *t* = 15, *Y*(15) = 27. (Time-2) 01/04/2020, *t* = 19, *Y*(19) = 41; (c) 01/21/2020, *t* = 36, *Y*(36) = 375; (Time-3) 01/22/2020, *t* = 37, *Y*(37) = 437; (Time-4) 01/23/2020, *t* = 38, *Y* (38) = 507; (Time-5) 01/24/2020, *t* = 39, *Y*(39) = 572; (Time-6) 01/25/2020, *t* = 40, *Y*(40) = 618. Note that the case data between 01/05/2020-01/20/2020 were discarded due to significant changes experienced in this time period in the case reporting requirements and practice.

Using the data described above, *R* is estimated to be 4.08, indicating that an infected patient infects more than four susceptible people during the outbreak. This value substantially exceeds WHO’s estimate of *R*_0_ (supposed to be smaller than *R*) between 1.4 and 2.5, and is also higher than a recent *R*_0_ estimate between 3.6 and 4.0 (*8*). Compared against the 2003 SARS epidemic, *R* of 2019-nCoV is higher than that of SARS in both Beijing (2.76) and Guangzhou (3.01) (calculated using the same method). To test the robustness of findings, we performed sensitivity analyses by adopting varying values of *L* and *D*, generated from a Gaussian distribution with *L*∼*N*(7,1) and *D*∼*N*(9,1). The resulting mean of R estimates is 4.08, as expected, with SD=0.36 (95% CI 3.37∼4.77).

To predict the future outbreak profile, we developed a model based on the deterministic Susceptible-Exposed-Infectious-Recovered-Death-Cumulative (SEIRDC) structure (*9*). Overall, our model appears to explain the reported case counts very well during the current early stage of the outbreak. An interesting finding is that by setting the start date to a time earlier than 12/16/2020 (the experimented range is from 12/01/2019—12/15/2019), the SEIRDC model is able to provide a better fit for the case counts. This indicates that human-to-human transmission may have started earlier than what the current prevailing viewpoint suggests. Obviously, further molecular and epidemiological studies are needed to draw any conclusions in this regard.

The SEIRDC model estimates the fatality rate for 2019-nCoV is 6.50%. As a base of comparison, the fatality rate for 2003 SARS was 7.66% and 3.61% for Beijing and Guangzhou, respectively. We used the model to predict the confirmed case counts and death counts in the first 80 days of the ongoing 2019-nCoV outbreak. We simulated these counts for the 2003 SARS outbreaks in Beijing and Guangzhou as well, using the case counts as input. The basic assumption is the absence of any control measures in all these scenarios. At the end of this 80-day period, according to our simulations, the 2019-nCoV case counts (35,454) is close to that of SARS in Guangzhou (37,663) and much higher than that of SARS in Beijing (17,594). The 2019-nCoV death count (1,089) is much higher than that of SARS in Guangzhou (725) and Beijing (690).

Our study also suggests that by reducing the average infectious period to <2.3 days, the resulting *R* will decease to a value less than 1, meaning the epidemic can be effectively controlled.

In conclusion, considering transmissibility and fatality rate, 2019-nCoV poses a major public health threat, at least at the level of 2003 SARS. Epidemiological studies are critically called for to evaluate the effectiveness of stringent measures such as lockdown and help the design of refinements and development of potential alternative strategies for the next phase of the 2019-nCoV outbreak.

## Data Availability

All data generated or analyzed during this study are included in this article.

## Acknowledgments

This work was supported in part by grants from the Ministry of Science and Technology (2016QY02D0305), National Natural Science Foundation of China (71621002, 71771213, 71790615, 71972164 and 91846301), Chinese Academy of Sciences (ZDRW-XH-2017-3), and the Hunan Science and Technology Plan Project (2017RS3040, 2018JJ1034).

## Disclaimers

Nil.

## Author Bio

Dr. Zhidong Cao is a researcher in the State Key Laboratory of Management and Control for Complex Systems, Chinese Academy of Sciences Institute of Automation. His primary research interests are infectious disease informatics, spatio-temporal data processing, and social computing.

## Footnotes

^1^ These first authors contributed equally to this article.

## Address for correspondence

Daniel Zeng, Chinese Academy of Sciences Institute of Automation, Room 1202, Automation Building, 95 Zhongguancun East Road, Haidian, Beijing 100190, China.

**Figure.**
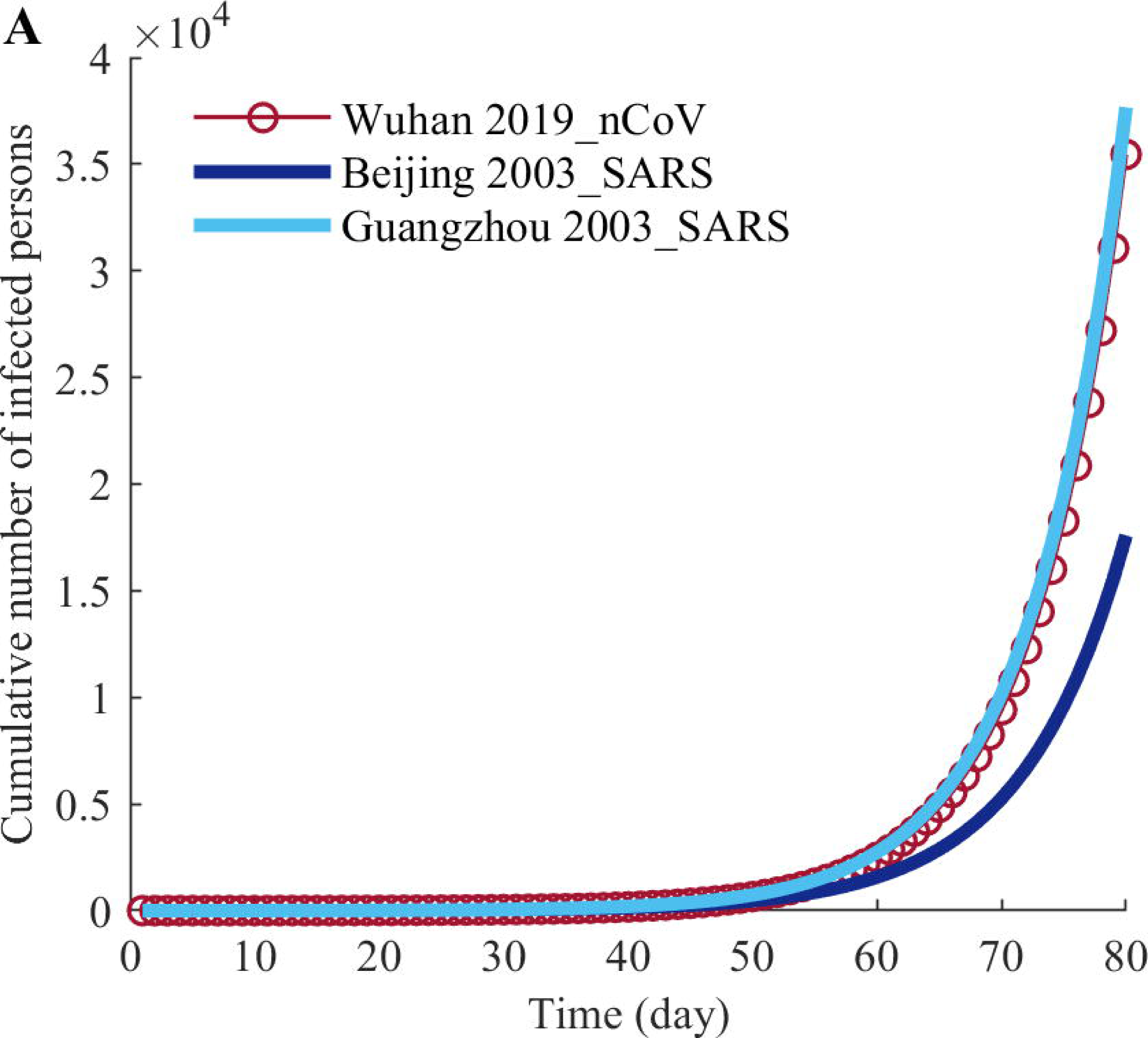
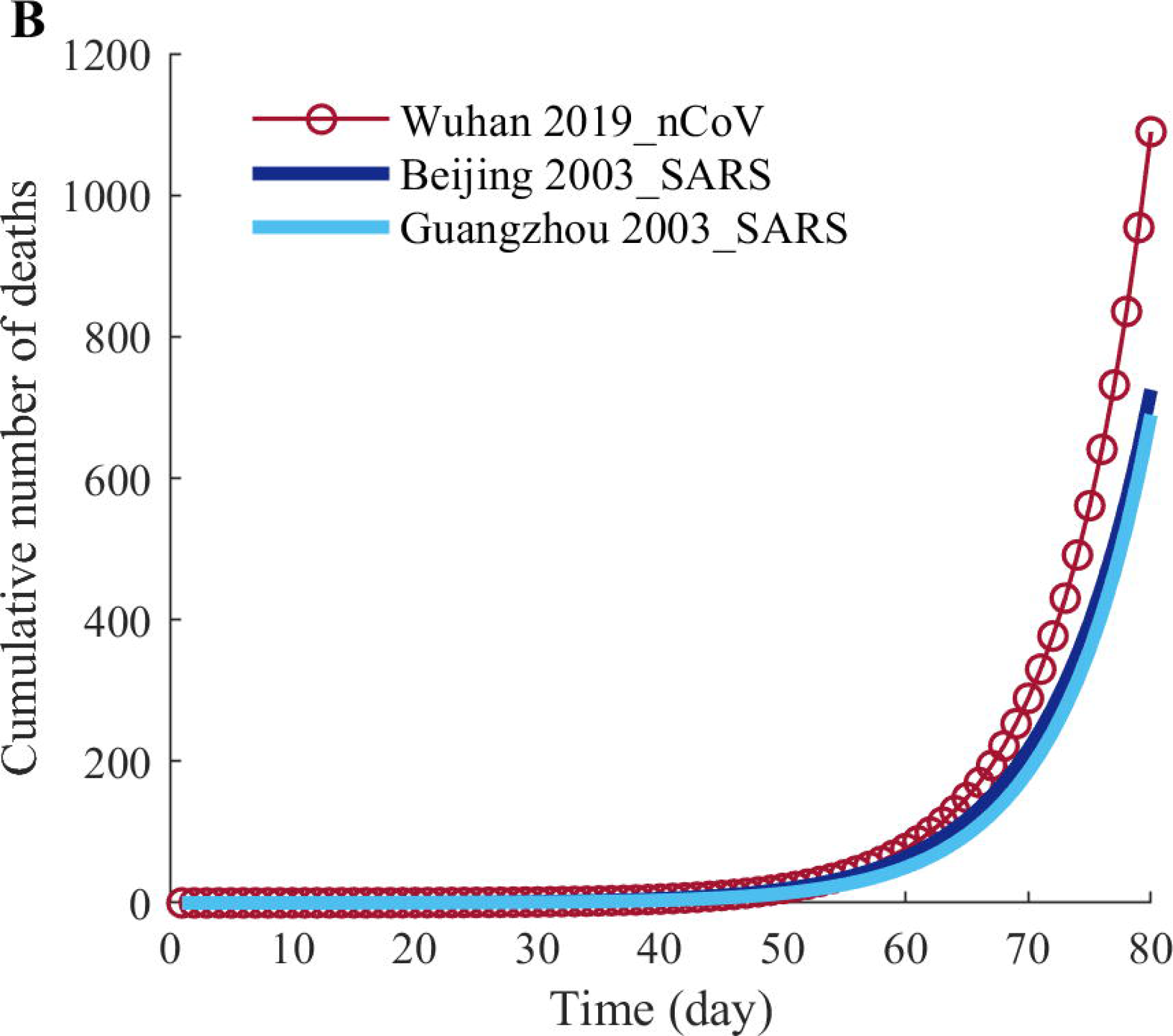
SEIRDC model predictions for (A) cumulative numbers of infected persons and (B) deaths of 2019-nCoV, 2003-SARS in Beijing, and 2003 SARS in Guangzhou in the first 80 days after the outbreak.

